# Importance of untested infectious individuals for the suppression of COVID-19 epidemics

**DOI:** 10.1101/2020.04.13.20064022

**Authors:** Francisco J. Pérez-Reche, Ken J. Forbes, Norval J. C. Strachan

## Abstract

The impact of the extent of testing infectious individuals on suppression of COVID-19 is illustrated from the early stages of outbreaks in Germany, the Hubei province of China, Italy, Spain and the UK. The predicted percentage of untested infected individuals depends on the specific outbreak but we found that they typically represent 50% to 80% of the infected individuals. Even when unreported cases are taken into account, we estimate that less than 8% of the population would have been exposed to SARS-CoV-2 by 09/04/2020 in the analysed outbreaks. These levels are far below the 70-85% needed to ensure herd immunity and would predict a resurgence of infection if ongoing lockdowns in the outbreaks are fully lifted. We propose that partially lifted lockdowns together with early and thorough testing allowing for quick isolation of both symptomatic and asymptomatic cases could lead to suppression of secondary waves of COVID-19 epidemics.

## Introduction

Daniel Defoe in “A Journal of the Plague Year” (1722) comments on the 1665 Great Plague of London that “… if all the infected persons were effectually shut in, no sound person could have been infected by them, because they could not have come near them. But the case was this (and I shall only touch it here): namely, that the infection was propagated insensibly, and by such persons as were not visibly infected, who neither knew whom they infected or who they were infected by.” ((*1*) COVID-19 is presenting similar problems today.

COVID-19 produced by the SARS-CoV-2 virus emerged in Wuhan, China in December 2019 (*2*). The virus has spread at an unprecedented rate since then, leading to 1,521,252 confirmed cases and 92,798 deaths distributed in 213 countries as of 10 April 2020 (*3*). The worldwide burden of the disease is still growing despite significant efforts in many countries to suppress the spread of the virus. So far, efforts have focused on non-pharmaceutical interventions which range from handwashing or social distancing to more stringent measures such as isolation of infected individuals, banning of large gatherings or severe lockdowns (*4, 5*).

Optimising interventions to mitigate or suppress the burden of COVID-19 remains a pressing global challenge due to significant uncertainties regarding the transmissibility of SARS-CoV-2 and other factors such as possibly a large proportion of undocumented infections as well as political, social and economic considerations (*6, 7*). Underreporting of infections may depend on the testing ability of different countries and the presence of asymptomatic infected individuals (*8*–*10*). There is no consensus on the proportion of unreported cases and their potential impact on the spread of SARS-CoV-2. For instance, a World Health Organization report in February suggested that “the proportion of truly asymptomatic infections is unclear but appears to be relatively rare and does not appear to be a major driver of transmission” (*11*). Studies testing for SARS-CoV-2 infection in both symptomatic and asymptomatic individuals (*8*–*10*), however, suggest that asymptomatic carriers can represent 50% or more of the cases. In many countries that mostly test individuals when they have symptoms, unreported infections are likely to include at least most of the asymptomatic individuals or those with mild symptoms. Such individuals can act as silent carriers for SARS-CoV-2 and have been suggested as a key factor promoting the rapid spread of the virus (*12*), similar to what has been observed in other infectious diseases (*13*). On the positive side, if recovery from infection leads to immunity, one could hope that untested positive individuals could significantly contribute to the build-up of herd immunity in the population (*14, 15*). It is not clear to what extent this could be the case. The importance of silent carriers on interventions for mitigation and suppression (*16*) of the infection is not clear either.

Mathematical modelling has been very successful in epidemiology (*17*–*19*) and there is an ongoing effort to propose models to describe the dynamics of COVID-19 epidemics (*4, 12, 15, 16, 20*–*26*). Unreported infectious individuals have been included in some models (*4, 12, 23, 27*) but their influence on control strategies has not been analysed.

Here, we use data from the outbreaks in Germany, Hubei (China), Italy, Spain and UK to calibrate a mathematical model that accounts for the force of infection associated with both tested (reported) and untested infectious individuals (see a brief description of the simplest version of the model in Figure 1 and more details in Methods). In order to compare outbreaks in different regions/countries, we fit the model independently to each outbreak. The calibrated model is used to study the effect of two suppression strategies: Interventions aiming for a reduction of transmission at the population level (representing, e.g., social distancing or a lockdown) and local interventions consisting in isolation of both tested and untested infectious individuals.

**Figure 1.**
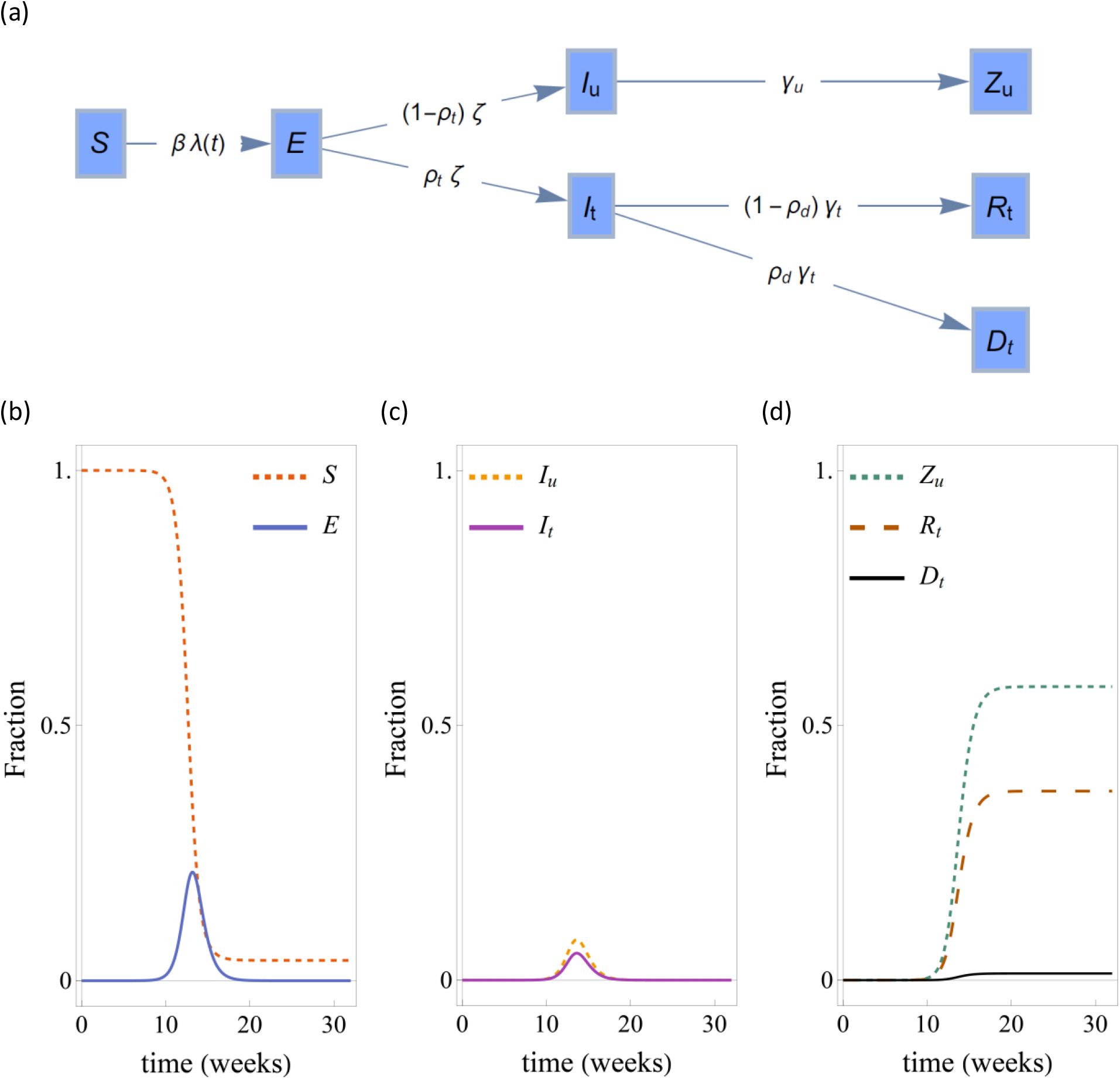
Simplified version of the compartmental model used to simulate the SARS-CoV-2 epidemic. (a) Flow diagram of the model. During epidemics, susceptible individuals (compartment *S*) become exposed to the virus (compartment *E*) at a rate *βλ*(*t*). Here, *β* is the rate at which an infected individual transmits infection to a susceptible individual and *λ*(*t*) is proportional to the number of infected individuals at time *t*. Exposed remain in this state during a latent period *ζ*^−1^ after which they become infectious. Of those that are infectious, a fraction *ρ*_*t*_ are tested for infection and move to the tested infected compartment *I*_*t*_. The remaining fraction of infectious individuals, 1 − *ρ*_*t*_, are not tested for infection and move to the untested infected compartment, *I*_*u*_, after the incubation period. A fraction *ρ*_*d*_ of infected individuals that were tested for infection, i.e. a fraction *ρ*_*d*_ of *I*_*t*_, die at a rate *γ*_*t*_ and move to compartment, *D*. The rest of tested infected move to the recovered compartment *R*_*t*_. Infected individuals in the untested compartment, *I*_*u*_, move to a compartment *Z*_*u*_ which contains individuals that were not tested for infection and recovered or died. Assuming that recovered individuals are fully immune to infection, individuals in compartment *Z*_*u*_ are effectively removed from the epidemic. Panels (b)-(d) show a typical time evolution of model variables. As the epidemic progresses, (b) the number of susceptible individuals decreases. The number of exposed individuals initially increases, reaches a peak and decreases at later stages of the epidemic. (c) The progression of the number of both tested and untested infected individuals also exhibits a peak. The decay of *I*_*t*_ and *I*_*u*_ after the peak induces a gradual weakening of the chain of transmission that leads to the end of the epidemic. (d) The number of tested and untested individuals that recover from infection or die increase monotonically during the epidemic.

### Model calibration, under-reporting and reporting delays

The model was calibrated using time series for the number of tested infected individuals and reported deaths (i.e. data for compartments *I*_*t*_ and *D* in the model of Figure 1). It is assumed that the number of deaths in the datasets originate from individuals that were tested for infection. Figure 2 shows that the model captures the trend of tested infected individuals and deaths at the early stages of the fitted outbreaks. Estimates for the parameters of each outbreak are given in Table 1.

**Table 1.** Estimates of the model parameters given in terms of the 5% percentile, median and 95% percentile. *β* is the transmission rate, *ρ*_*t*_ is the proportion of tested infectious (in percentage), *ρ*_*d*_ is the proportion of tested infectious that die (in percentage), *γ*_*t*_ is the rate of recovery of tested infectious individuals, *γ*_*u*_ is the rate of recovery of untested infectious individuals, *E*(0) is the initial number of exposed individuals and ℛ_0_ is the reproduction number.

| | $\beta$ (day<sup>-1</sup>) | $\rho_t$ (%) | $\rho_d$ (%) | $\gamma_t$ (day<sup>-1</sup>) | $\gamma_u$ (day<sup>-1</sup>) | $E(0)$ | $\mathcal{R}_0$ |
| --- | --- | --- | --- | --- | --- | --- | --- |
| Germany | (0.6,0.8,1.0) | (31,53,87) | (0.4,1.7,3.1) | (0.05,0.15,0.24) | (0.12,0.27,0.42) | (544,3976,6939) | (2.0,3.2,6.4) |
| Hubei | (1.0,1.2,1.4) | (22,40,57) | (2.0,3.3,5.0) | (0.14,0.24,0.36) | (0.18,0.31,0.39) | (1626,3101,6526) | (3.2,3.9,5.8) |
| Italy | (0.9,1.2,1.4) | (10,25,50) | (2.7,3.7,4.9) | (0.21,0.30,0.38) | (0.17,0.31,0.38) | (1165,2271,5302) | (3.2,3.9,5.9) |
| Spain | (1.0,1.3,1.4) | (19,33,62) | (2.7,3.7,4.8) | (0.23,0.32,0.38) | (0.18,0.30,0.38) | (1207,2620,5590) | (3.6,4.2,6.2) |
| UK | (0.8,1.0,1.3) | (9,16,31) | (2.2,3.2,4.9) | (0.17,0.28,0.39) | (0.13,0.26,0.38) | (1031,2452,5179) | (3.2,4.2,5.8) |

**Figure 2.**
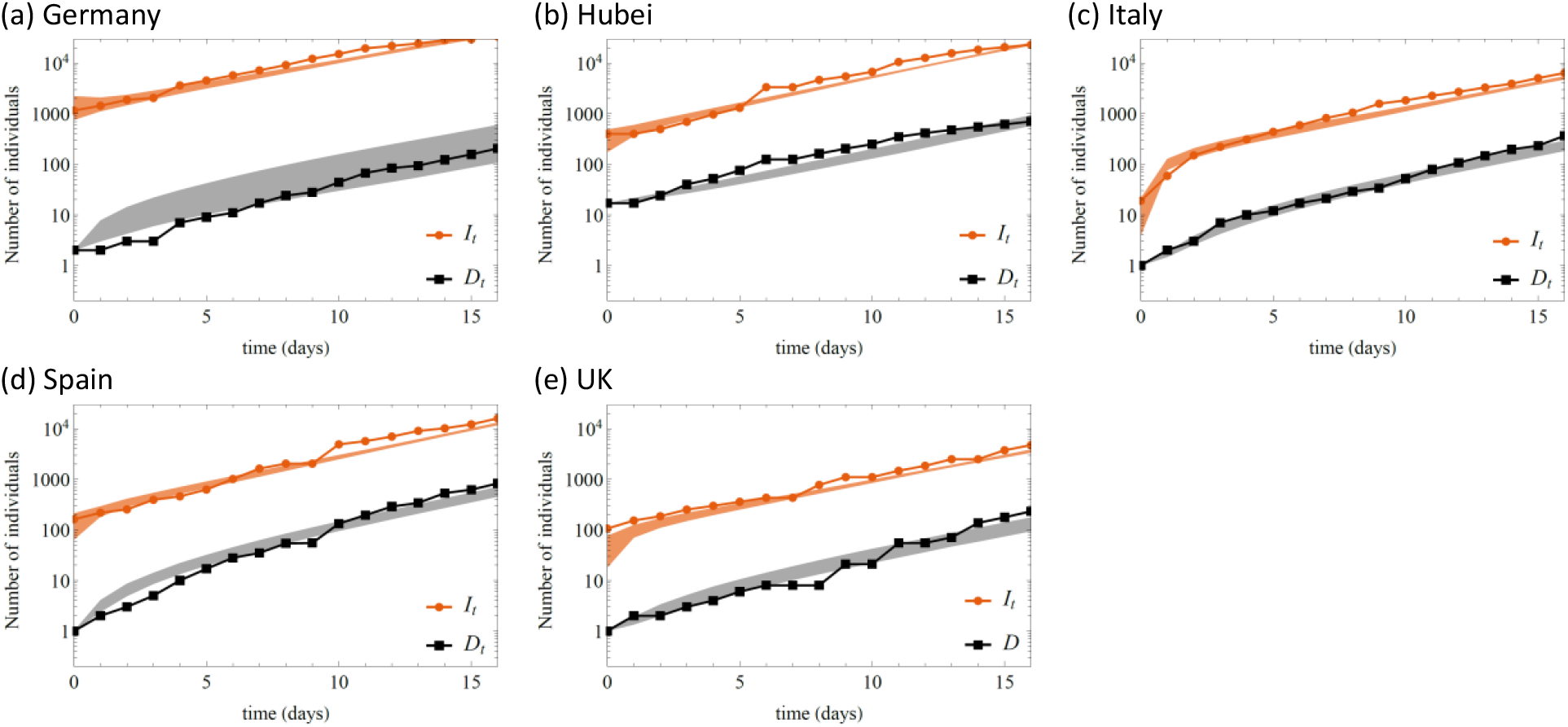
Number of tested infected (*I*_*t*_, red) and dead (*D*_*t*_ black/grey) individuals registered in (a) Germany, (b) Hubei, (c) Italy, (d) Spain and (e) UK. Symbols show the data and shaded regions show the 90% confidence interval of model predictions at any given time. Time given in days since the first day with a positive number of deaths in the datasets (see Table 2). Logarithmic scale is used in the vertical axis of each plot.

The obtained values for the percentage of infected individuals that were tested, *ρ*_*t*_, reveal that during the early stage of outbreaks, Germany scored the highest in terms of testing for infection (median 53%). Hubei follows Germany in terms of testing, followed by Spain, Italy and the UK. Our prediction for Hubei is not far from the 65% reporting percentage estimated by Li et al. (*12*) for China in the period considered here. The high testing percentage predicted for Germany agrees with the known high testing capacity in this country (*28*). Taking the confidence intervals into account, we estimate that for each infected individual tested in the UK, there could have been between 2 and 10 untested infected individuals. At the other end of the testing spectrum, we estimate that for each infected individual tested in Germany, between 0.2 and 2 individuals might have not been tested at the beginning of the epidemic. A higher testing percentage for Germany is in qualitative agreement with estimates given elsewhere (*23, 27*). Our estimates for the reporting percentage, however, tend to be higher than those obtained by Jagodnik et al. (*27*) and the differences we found between countries are not as extreme as those given by Chicchi et al. (*23*)

**Table 2.** Details on the first day used to calibrate the models, number of deaths by that day, date when a lockdown was ordered in each of the countries/region analysed, and population of each country/region.

| Country/Region | First day considered | Number of deaths on first day considered | Date when lockdown ordered(4, 5) | Population, $N$ |
| --- | --- | --- | --- | --- |
| Germany | 9/3/2020 | 2 | 22/03/2020 | 82,114,224 |
| Hubei | 22/1/2020 | 17 | Wuhan 23/1/2020,<br>Hubei province<br>13/2/2020 | 58,160,000 |
| Italy | 21/2/2020 | 1 | 11/3/2020 | 59,359,900 |
| Spain | 3/3/2020 | 1 | 14/3/2020 | 46,354,321 |
| United Kingdom (UK) | 5/3/2020 | 1 | 24/3/2020 | 66,181,585 |

Assuming that the testing percentage for infection remains constant during the course of epidemics and no control interventions are implemented, our model predicts that the number of tested and untested infected individuals would evolve in parallel in all the studied outbreaks which would last for around 12 weeks in all cases (see Figure 3). We see, however, that the epidemic in Germany would be different in the sense that the number of untested infected individuals remains smaller than the number of tested individuals during the whole epidemic. Italy, Spain and UK exhibit the opposite behaviour with more untested than tested individuals. For Hubei, we predict similar levels of untested and tested percentages. Obviously, these predictions will not be fulfilled since control interventions are imposed in all these countries and testing strategies might change during the pandemic.

**Figure 3.**
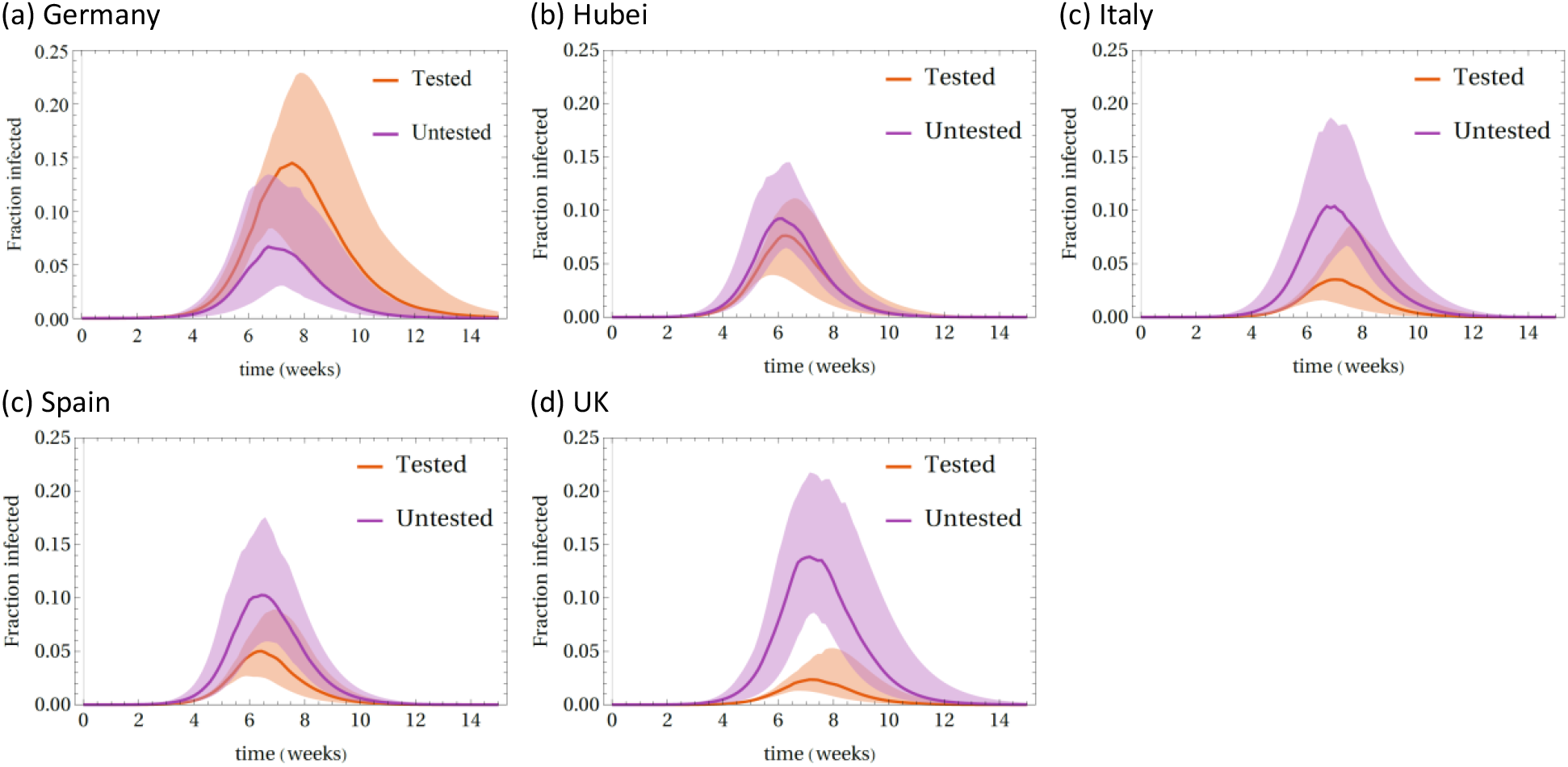
Fraction of the susceptible population in (a) Germany, (b) Hubei, (c) Italy, (d) Spain and (e) UK of tested (*I*_*t*_, red) and untested (*I*_*u*_, purple) that are infected (assuming that no control interventions are implemented during the course of epidemics). Lines give the median of the model predictions and shaded areas give the 90% confidence interval of predictions at any given time. Time given in weeks since the first day considered for each country/region (see Table 2 in Methods).

From our estimates of *ρ*_*t*_, we predict that 47% [90% CI:13% - 69%] of infected individuals were not tested for infection in Germany. Bearing in mind that 50% of infected individuals might be asymptomatic (*8, 10*), we conclude that most of those that were infected but not tested in Germany were asymptomatic. In contrast, the higher percentages of untested infected individuals predicted for other countries suggest that untested individuals in such countries might include a significant number of individuals with symptoms in addition to those that are asymptomatic. In particular, these may include infections of care home residents that are known to be underreported in many countries (*29*).

The proportion of tested infected individuals that die, *ρ*_*d*_, is smaller for the outbreak in Germany than for the other outbreaks. This might be a combined effect of the fact that infected individuals in this country were relatively young at the beginning of the outbreak (*30*) and the high testing rate. Indeed, the COVID-19 fatality rate is lower for the younger than for the elderly (*31*) and, the higher the testing rate, the more individuals with mild or no symptoms will be included in the tested infected compartment of our model. The lower death rate of individuals with mild symptoms will lead to an effectively lower death rate for the whole set of infected individuals in this compartment. Accordingly, a lower value of *ρ*_*d*_ does not necessarily mean a lower overall infection fatality rate (i.e. proportion of the population that dies). In fact, we found that the predicted fraction of deaths by the end of unmitigated epidemics is not too different for different countries (medians are as follows: 0.4% for Germany, 0.9% for Hubei, 0.5% for Italy, 0.8% for Spain and 0.3% for UK). We remark that these percentages correspond to deaths of infected individuals that were tested for infection since these are the deaths captured by our model (see Figure 1). Deaths of untested infected individuals such as unreported from care homes (*29*) are simulated by our model in terms of the compartment *Z*_*u*_ which does not explicitly separate deceased individuals from those that recovered from infection.

The median of the recovery rate from infection, *γ*_*t*_, gives the following estimates for the time 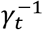 from reporting of infection to recovery or death: 3 days for Spain, 3.3 days for Italy, 3.6 days for the UK, 4.2 days for Hubei and 7 days for Germany. The time for Hubei is consistent with the 3.48 days reported by Li et al. (*12*) for China. In general, the values we obtained are smaller than the infectious period (time from infection to death or recovery) reported elsewhere for COVID-19 (*4, 31, 32*). Our estimates thus probably reflect a reporting delay in all the studied outbreaks, in agreement with data on the onset of symptoms and reporting (*30, 33, 34*). Our model predicts the smallest reporting delay for Germany. This is again in agreement with the high testing capacity of this country.

The removal period for untested infected individuals, 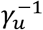, takes values of around 3 days for all the studied outbreaks. Comparing with the reporting-to-recovery period 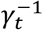and bearing in mind the reporting delays in all outbreaks, our estimates of 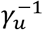 suggest that untested individuals remain infectious for a shorter time than tested individuals. This is in line with the lower infectivity of untested individuals proposed in a recent study that, instead of assuming different recovery rates for tested and untested individuals, assumed a lower transmission rate for unreported infectious individuals (*12*).

We predict that the number of exposed individuals at the beginning of our simulations, *E*(0), is of the order of several thousand for all the countries, in qualitative agreement with estimates of a previous study for China (*12*).

We obtained similar values of the reproduction number ℛ_0_ for all the studied outbreaks. To some extent this reflects our prior assumption that transmission of SARS-CoV-2 is intrinsically similar in different regions. The transmission rate, *β*, was derived from the estimates of *ρ*_*t*_, *γ*_*t*_, *γ*_*u*_ and ℛ_0_, using Eq. (4) (see Methods). Values of *β* are statistically similar for all countries except Germany which features a smaller value. Bearing in mind that ℛ_0_ and *γ*_*u*_ take similar values for all the countries, we conclude that a lower value of *β* is a consequence of the higher testing rate and smaller recovery rate *γ*_*t*_ for this country. Indeed, according to Eq. (4), *β* decreases with increasing *ρ*_*t*_ and decreasing *γ*_*t*_ (or increasing period 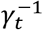).

### Interventions focusing on the reduction of the transmission rate at the population level

Interventions such as lockdowns or social distancing can be effectively studied by reducing the transmission rate *β* in our model. As illustrated in Figure 4(a), the outbreak can be significantly delayed if the transmission rate is reduced from early stages in the epidemic, in agreement with other works (*16*). In spite of that, the predicted number of deaths by the end of the epidemic only reduces significantly when *β* is reduced by a factor close to *r* = 1 − 1/ℛ_0_ to ensure an early eradication of the infection (*17*). Based on our estimated values for ℛ_0_, this requires reducing the transmission rate by more than 70% in all of the studied outbreaks. This is illustrated in Figure 4(b) for the UK. As can be seen, the number of deaths would only reduce significantly if the number of contacts were reduced by approximately 80%.

**Figure 4.**
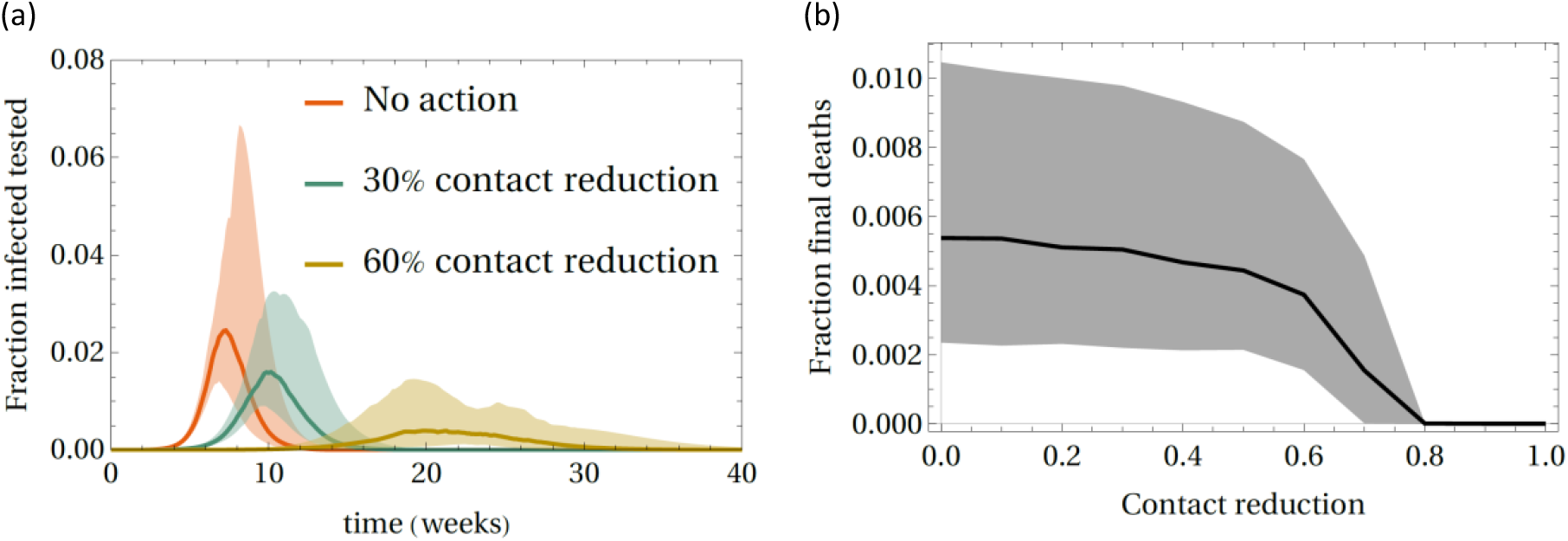
Predicted effect of reducing the transmission rate *β* on the outbreak in the UK in a hypothetical scenario in which the intervention was applied from the beginning of the epidemic and kept active until the end. (a) Proportion of tested infectious individuals, *I*_*t*_, as a function of time if no interventions are implemented (red) or if the transmission rate is reduced by 30% (green) or 60% (brown). (b) Effect of transmission reduction on the fraction of the susceptible population that die, *D*_*t*_, during the epidemic. The line gives the median and the shaded regions give the 90% confidence interval of predictions at any value of the reduction of the transmission rate.

Our model can be readily used to predict the effect of lockdowns of arbitrary duration and different exit strategies from such lockdowns.

Figure 5 shows predictions for the lockdowns in Hubei, Italy and Spain. Predictions for Germany and the UK are also possible but a comparison with observed effects is uncertain at present since lockdowns were implemented more recently in these countries.

**Figure 5.**
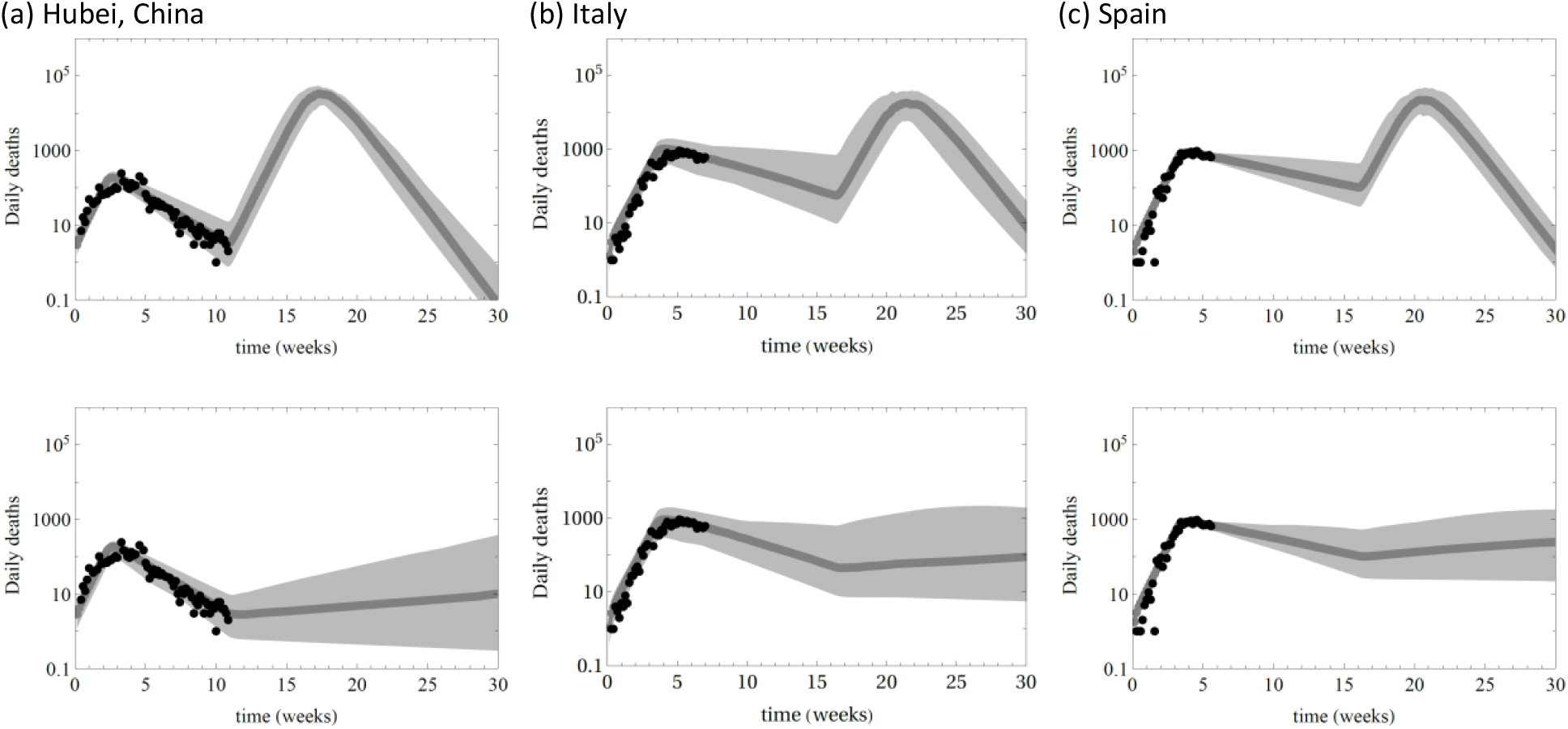
Prediction for the daily deaths,*D*_*t*_, during and after lockdowns in Hubei, Italy and Spain. (a) Upper panel: Predicted effect of a lockdown in Hubei with a 90% reduction of transmission for 60 days (∼8.5 weeks) and return to full transmission after the lockdown. Lower panel: A 60 days lockdown as in the upper panel followed by a weaker lockdown with 75% reduction of transmission. (b) Upper panel: Predicted effect of a lockdown in Italy with transmission reduced by 80% for 90 days (∼13 weeks) and return to full transmission after the lockdown. Lower panel: A 90 days lockdown as in the upper panel followed by a weaker lockdown with 70% reduction of transmission. (c) Upper panel: Predicted effect of a lockdown in Spain with transmission reduced by 80% for 90 days and return to full transmission after the lockdown. Lower panel: A 90 days lockdown as in the upper panel followed by a weaker lockdown with 72% reduction of transmission. Symbols represent data, the thick grey line gives the median of the predictions and shaded areas give the 90% confidence interval. Logarithmic scale is used in the vertical axis of all the plots.

For Hubei, the model reproduces well the observed daily deaths assuming a 90% reduction of the transmission rate from 6 February 2020 (see Figure 5(a)). The later is an effective date between the 23 January when the lockdown was implemented in Wuhan, the capital of the province, and 13 February when it was implemented in the whole province. Despite the spectacular reduction of daily deaths induced by the lockdown in Hubei, our model predicts that fully removing the lockdown after 60 days (i.e. approximately at the time of writing) would lead to a rapid resurge of the epidemic (see the marked increase predicted after week 12 in Figure 5(a)). In contrast, an exit strategy in which the transmission is kept reduced by a 75% is predicted to keep the number of daily deaths below 100 for many weeks (see Figure 5(a)).

We estimate that the lockdown ordered in Italy reduced the transmission by around 80%. Our prediction suggests that this will lead to a significant decrease of the number of deaths if it is kept for a long enough time. In particular, Figure 5(b) shows a scenario in which the lockdown is kept at the same level for 90 days since its implementation on 11 March 2020. In this case, we predict around 69 [90% CI: (16,568)] daily deaths at the end of the lockdown. As for Hubei, a full removal of the lockdown leads to a fast resurge of the epidemic (Figure 5(b)). For Italy, we estimate that an exit strategy from this lockdown should still keep the transmission at low values (∼70% reduction) for the daily deaths to remain at a moderate value of around 100 (see Figure 5(b)).

The effectiveness of the lockdown imposed in Spain is predicted to be similar to the one in Italy (Figure 5(c)): The current lockdown managed to reduce the transmission by ∼80% and resurge of infection is predicted to occur if the lockdown is completely removed after 90 days. At the end of the initial 90 days lockdown, we predict around 99 [90% CI: (22,501)] daily deaths. The number of daily deaths is predicted to remain at this level if the lockdown is partially lifted to a situation in which transmission is kept to a 72% reduced level.

Irrespective of the effectiveness of the lockdown, our model predicts that epidemics will resurge after relatively extended lockdowns in Hubei, Italy and Spain. The same is likely to occur in other countries. In fact, resurgence of the disease is predicted even for much longer lockdowns. This is due to the fact that an early lockdown delays the spread but does not lead to herd immunity. Assuming that recovered individuals are immune to SARS-CoV-2, herd immunity is only achieved when a proportion 1 − 1/ℛ_0_ of the susceptible population has been infected and died or recovered. Even if the number of untested individuals that may have recovered are taken into account, we estimate that, as of 09/04/2020, the proportion of the susceptible population that has been exposed to the virus (i.e. the attack rate, see Methods) is 0.65% [90% CI: 0.5%-1.2%] for Germany, 0.5% [0.3%-1.0%] for Hubei, 4.6% [3.3%-7.2%] for Italy, 3.7% [2.0%-6.4%] for Spain and 4.4% [2.7%-6.9%] for the UK. These proportions are small compared to the 70-85% needed to ensure herd immunity for these epidemics. Our conclusion is in qualitative agreement with the results by Flaxman et al. (*4*) despite the fact that their estimates for the attack rate tend to be higher than ours. Our conclusions, however, disagree with Lourenço et al. (*15*) that predicted much higher attack rates that would be close to herd immunity.

### Isolation of infected individuals

Prompt isolation of infected individuals is regarded as an effective strategy to reduce the transmission of infection and significantly reduce the size of epidemics (*35*). The presence of silent infectious carriers, however, makes the implementation of this strategy challenging for SARS-CoV-2.

In order to study the effect of isolating tested and untested individuals, we extended the model shown in Figure 1 to include compartments for isolated tested and untested infected individuals (see Model 2 in Methods). Interventions are parametrised by the fraction of tested (untested) infected individuals that are randomly selected to be isolated, *ρ*_*Qt*_ (*ρ*_*Qu*_), and the rate *δ* at which they are isolated after testing/reporting. Isolation strategies leading to eradication of infection satisfy the condition ℛ_0*Q*_ < 1, where ℛ_0*Q*_ is the reproduction number for Model 2 (see Eq. (7) in Methods). Given the significant reporting delays we found, isolation has to be fast after testing positive for eradication of infection to be possible. In particular, we found that isolation after an average time of *δ*^−1^= 1 day could only lead to eradication in Germany; for other countries, ℛ_0*Q*_ remains larger than 1 for any value of *ρ*_*Qt*_ and *ρ*_*Qu*_. Following this, in Figure 6 we show results for a scenario in which individuals are isolated after an average time of *δ*^−1^= 0.5 days. In this case, eradication is possible in all the studied countries if *both ρ*_*Qt*_ and *ρ*_*Qu*_ are large enough, i.e. if enough tested and untested cases are isolated. The estimated boundaries separating the eradication region (ℛ_0*Q*_ < 1) from the epidemic region (ℛ_0*Q*_ > 1) are different for different countries but differences are statistically less marked if confidence intervals are taken into account.

**Figure 6.**
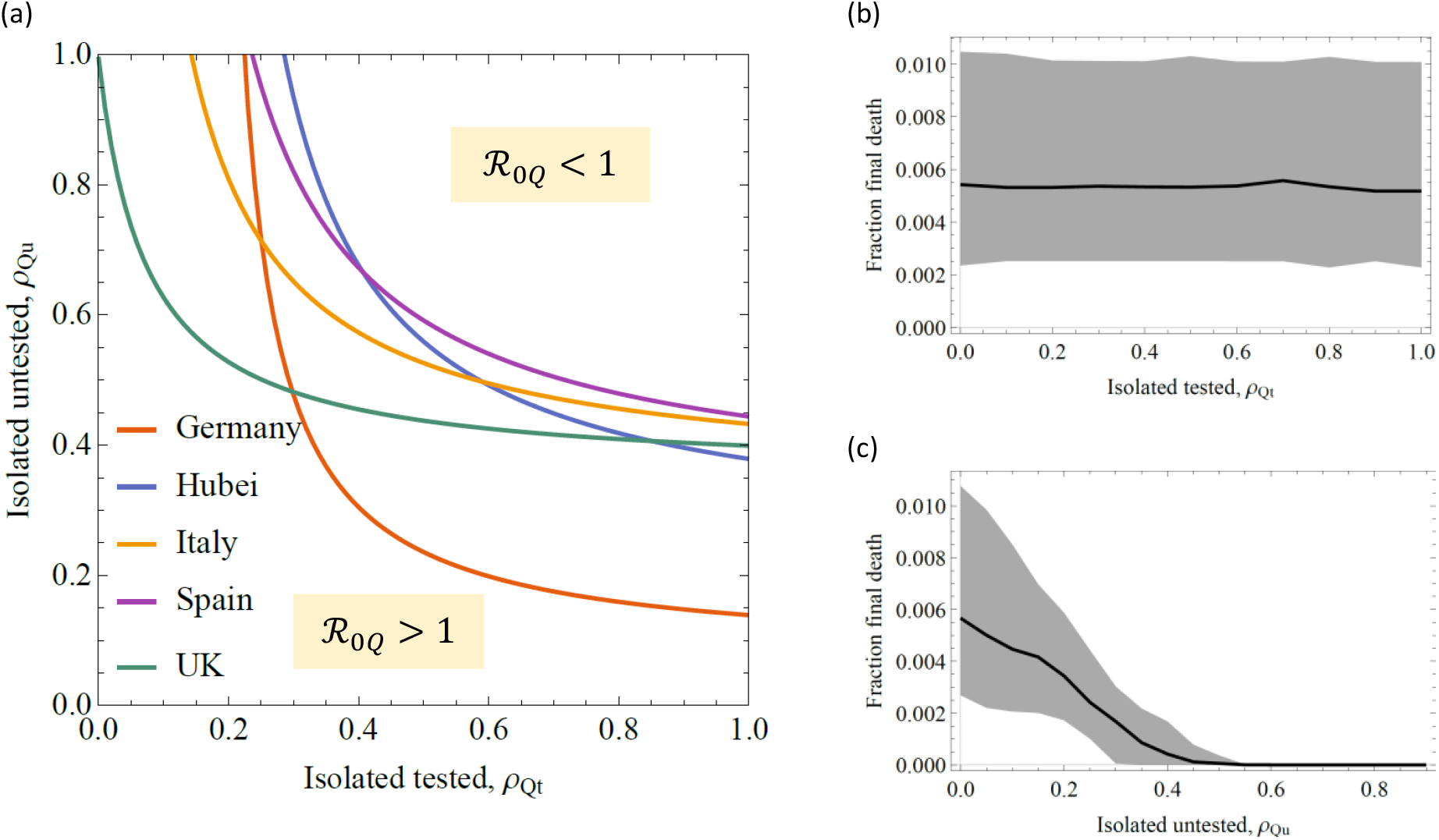
Interventions in which a fraction *ρ*_*Qt*_ of tested infected individuals and a fraction *ρ*_*Qu*_ of untested infected individuals are isolated in an average time of *δ*^−1^ = 0.5 days after testing. (a) Lines separating the regions in the space (*ρ*_*Qt*_, *ρ*_*Qu*_) where eradication occurs (above the line for a given country) from the regions where the epidemic grows (below the line). The lines are based on the median of ℛ_0*Q*_; confidence intervals are not shown for clarity. (b) Interventions that isolate a fraction *ρ*_*Qt*_ of tested infected individuals but do not isolate any untested infectious individual (i.e. interventions with *ρ*_*Qu*_ = 0). Parameters for the outbreak in the UK are used as an example. The line gives the median of the predicted fraction of susceptible individuals that die, *D*_*t*_, at the end of epidemics as a function of *ρ*_*Qt*_. (c) Interventions in which 70% of tested infected individuals are isolated (*ρ*_*Qt*_ = 0.7). The line gives the median of the predicted fraction of susceptible individuals that die at the end of epidemics as a function of the proportion *ρ*_*Qu*_ of untested individuals isolated. The shaded regions in (a) and (b) give the 90% confidence interval for the predicted fraction of deaths.

Interventions that only isolate tested infected individuals (i.e. with *ρ*_*Qu*_ = 0) are predicted to have a minor effect on the final fraction of deaths even if they manage to isolate all tested individuals (see Figure 6(b)). Isolation of tested individuals is not effective due to the underlying transmission associated with silent carriers that are not isolated and keep ℛ_0*Q*_ > 1.

Successful eradication of infection requires isolating both tested and untested infected individuals. For instance, an intervention in the UK in which 70% of tested infected individuals were isolated in *δ* = 0.5 days could drastically reduce the number of deaths if 40% of untested infected were isolated at the same rate. For Germany, the percentage of untested individuals that should be isolated to ensure eradication in this situation is around 15% since testing seems to be already faster and more effective than in other countries. Isolating infected individuals within half a day of being sampled is likely to be difficult to implement in practice. In order to allow for a longer time between reporting of infection and isolation, carriers of the virus should be identified earlier than currently done in most countries. This highlights the importance of fast identification of infected individuals. Identification of asymptomatic cases is expected to be challenging. However, we believe that efficient tracing of the contacts of symptomatic individuals and early testing of such contacts could facilitate the identification of asymptomatic cases.

### Implications for policy and conclusions

The main aim of our models is to contribute to the understanding of the epidemiological patterns of SARS-Cov-2 rather than to provide exact predictions. Hence the models should be viewed as a general guide of how the outbreak and interventions may play out rather than as an exact representation of COVID-19 epidemics. In spite of our simplifying assumptions, there are two main implications from the models which are directly relevant for policy in dealing with the outbreak.

The first, involves the existence of a significant proportion of cases that are not tested and may act as silent carriers of the infection. We found that the predicted percentage of untested infected individuals may represent 50% to 80% of the cases in Germany, Hubei, Italy, Spain and the UK. The specific percentage depends on the country and we found the lowest proportion of unreported cases in Germany. Based on studies in Iceland (*10*) and the Diamond Princess cruise (*8*), we conclude that asymptomatic infected individuals are likely to be the main contribution to the untested cases in all analysed outbreaks but a fraction of cases with mild symptoms are also likely to be untested. Even when unreported cases are taken into account, we estimate that less than 8% of the population would have been exposed to SARS-CoV-2 by 09/04/2020 in the analysed outbreaks. In policy terms, our results demonstrate that the current suppression strategies being employed in Germany, Hubei, Italy, Spain and the UK will not facilitate sufficient levels of herd immunity in the population that would control and eventually eradicate the virus. This leaves the risk of re-emergence of the virus once suppression strategies are lifted, similar to second waves of infection observed in the 1918 influenza epidemics (*36*). We predict, however, that partial relaxation of ongoing lockdowns could keep the number of daily deaths to less than 100.

The second implication involves the finding that unreported cases play an important role in the control of COVID-19 epidemics. In particular, unreported cases act as silent carriers and control strategies would need to account for them or be prone to the risk of re-emergence or ineffective suppression of spread. For instance, we predict that isolation of infected individuals can have a limited impact on the suppression of spread unless it includes silent carriers that are currently missed by most countries. In line with previous suggestions (*15, 37*), we suggest that thorough testing combined with contact tracing (*21, 22*), isolation of infected individuals and social distancing can be more effective to suppress SARS-CoV-2 spread than severe lockdowns. An option that was not available in the time of Daniel Defoe. However, in “A Journal of the Plague Year” (1722), Daniel Defoe wrote “…the shutting up of houses was a subject of great discontent, […] and the complaints of the people so confined were very grievous.” in relation to the lockdown imposed to combat the 1665 Great Plague of London (*1*). Similar feelings may apply to lockdowns ordered to combat COVID-19. At present, however, lockdowns are probably the most effective way to delay epidemics until effective pharmaceutical (e.g. vaccine, antivirals) or non-pharmaceutical interventions (e.g. early and thorough testing) become feasible.

## Materials and Methods

### Data

Data on numbers of infected and deceased individuals by country or region were obtained from the Wolfram Data Repository (*38*). Models were calibrated by considering data from the first available day in which the number of deaths is non-zero, as listed in Table 2. The date when lockdowns were ordered in each of the countries/regions is also given in Table 2.

### Models

We used extensions of the SEIR model (*18*) to include two types of infected individuals described by the compartments *I*_*t*_ and *I*_*u*_ (see Figure 1). The SEIR model with a single compartment for infectious individuals has already been used to describe the COVID-19 outbreak in China (*20, 24*) and a model with two compartments for infected individuals analogous to those proposed here was used by Li et al. (*12*)

#### Model 1

The model shown in Figure 1 is run with deterministic, continuous-time dynamics given by the following differential equations:

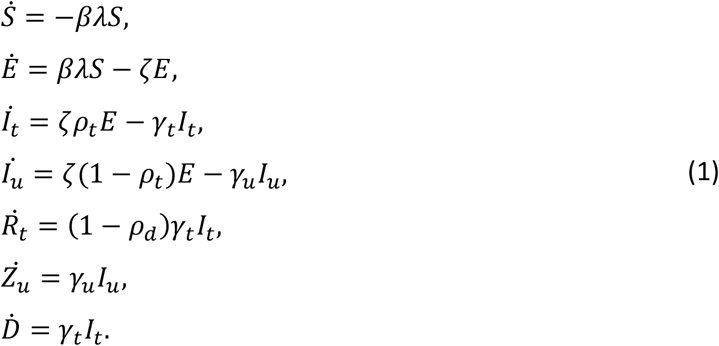

The force of infection in this model is *βλ*, where *β* is the transmission rate and

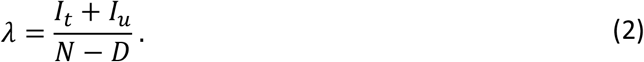

Here, *N* is the population size.

The attack rate at a given time *t* is defined as the fraction of individuals in a population of size *N* that have been exposed to the disease by that time. For the model described by Eqs. (1), we calculate the attack rate as follows:

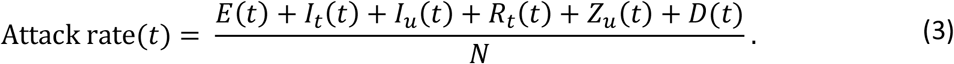

The reproductive number corresponding to this model can be analytically calculated using the next generation method (*19*) and is given by

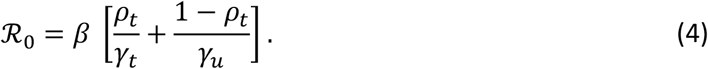

#### Model 2 – Isolation of infectious individuals

Isolation of infectious individuals is modelled by adding two more compartments, *Q*_*t*_and *Q*_*u*_, which contain isolated tested and untested infectious individuals, respectively (see Figure 7). The fraction of tested and untested infectious individuals are denoted as *ρ*_*Qt*_ and *ρ*_*Qu*_, respectively. Both types of infectious individuals are assumed to become isolated at the same rate, *δ*. The number of individuals in each compartment evolve according to the following differential equation:

**Figure 7.**
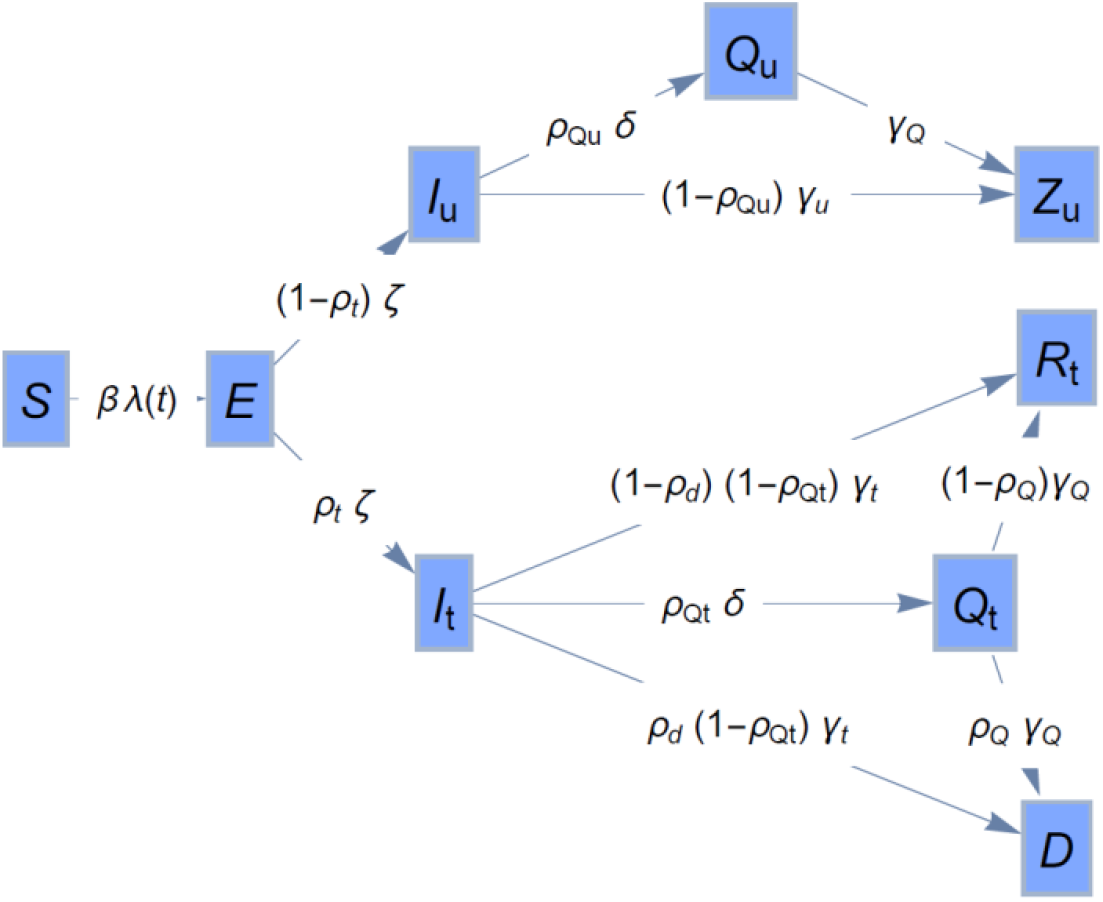
Extension of the basic epidemic model of Figure 1 to incorporate a compartment *Q*_*t*_ for isolation of a tested infectious individual and a compartment *Q*_*u*_ for isolation of untested infectious individuals.

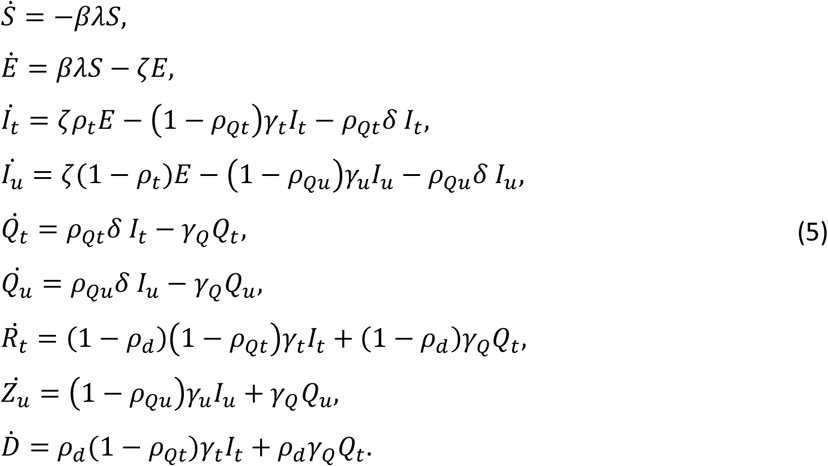

Here,

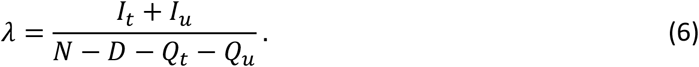

One can again use the next generation method (*19*) to obtain the following expression for the reproduction number:

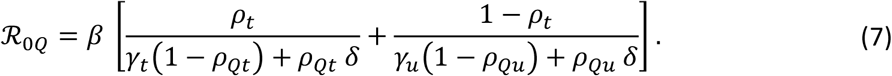

#### Main assumptions of the models

Several simplifying assumptions were made to keep a reasonable level of transparency in our models to enhance understanding of the analysed epidemics.

##### Homogeneous populations

The total number of individuals in each compartment of our models represent average values for the whole country. More precise descriptions at smaller scales such as geographical regions within countries or at the level of individuals would require accounting for spatial heterogeneity within the populations (*26, 39*) (e.g. cities and rural areas) as well as differences in both susceptibility and mortality across different age and vulnerability groups (*31, 40*) or topological heterogeneity of the network of contacts between individuals (*41*).

Ignoring heterogeneities limits the ability of our models to identify specific ways to make interventions operational. For instance, reductions in transmission are treated at a generic level without specifying if they could be achieved by enhanced social distancing, school closure, etc. Accounting for such details would require using individual-based simulations (*16*).

##### Deterministic dynamics

We focused on stages of epidemics in which the number of infectious individuals is large enough as for stochastic effects to be relatively unimportant on average (*22*). Our models can be extended to incorporate stochasticity (*42, 43*). This would give a more accurate description of epidemics when SARS-CoV-2 has just invaded or at later stages when the number of infected individuals becomes very low.

##### Imported cases

We focused on epidemics that are at a stage in which imported cases are expected to play a secondary role relative to internal transmission. Accounting for imported cases is crucial, however, to prevent re-emergence of infection once an area has reached a low numbers of infected individuals (*44, 45*). Imported cases could be included in our model in terms of an influx of infected individuals. In scenarios in which imported cases represent an important fraction of new infections, however, a stochastic version of the model would be more appropriate than the deterministic dynamics used here.

##### The transition times between compartments are exponentially distributed

This memory-less assumption is usual for classical compartmental models (*18*). For COVID-19, however, transitions between compartments are better described in terms of gamma distributions (*4, 31*) and using models with memory would provide a more precise description of the dynamics (*4, 18, 46*).

##### Immunity follows after recovery from infection

Whether or not this is the case, and if it is the duration of the immunity is still unclear (*47*). Our model could easily be extended to account for re-infections, should such data become available, and predictions might significantly change.

##### The latent and incubation periods coincide

We adopted a parsimonious approach which assumes that the latent period (i.e. the time between exposure to communicability) coincides with the incubation period (time between exposure and the appearance of symptoms) (*20*). There is, however, a growing body of evidence for pre-symptomatic transmission (*48*–*52*). There is the potential to incorporate this in our models by varying the incubation rate parameter, *ζ*, or including a new compartment for pre-symptomatic infectious individuals (*26*). We expect our predictions to be qualitatively independent of these details.

##### All of the population at the start of the epidemic are susceptible

However, it may be that a proportion are not for genetic reasons (*53*) or due to cross immunity (*54*).

### Parameter estimation

We fitted the Model 1 to data. Values for the incubation rate was set to (*40*) *ζ* = 1/5.2 days^-1^. The free parameters in our fits were the rate of transmission, *β*, proportion of infectious that were tested, *ρ*_*t*_, proportion of tested infectious that die, *ρ*_*d*_, rate to recovery of tested infectious individuals, *γ*_*t*_, rate of recovery of untested infectious individuals, *γ*_*u*_, and initial number of exposed individuals, *E*(0). We denote the free parameters by a vector *θ* = {*β, ρ*_*t*_, *ρ*_*d*_, *γ*_*t*_, *γ*_*u*_, *E*(0)}. The model was fit to the time series for the number of daily reported infected individuals and cumulative deaths, 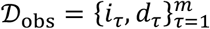, in a period of *m* days in the early stages of epidemics (here, *τ* is used to denote discrete time in days). In particular, we used *m* = 15 days since the first data point with a positive number of deaths (see Table 2). We used data at early stage of each outbreak to minimise the influence of suppression strategies on our parameter estimates. Using data on deaths is important to obtain reliable descriptions of COVID-19 epidemics since data on deaths is likely to be more accurately recorded than data on infected and recovered individuals (*4, 15, 27, 55*). In addition to deaths, we can use data on infected individuals which is represented by the tested infectious compartment, *I*_*t*_, in our models.

Our fitting procedure aims at calculating the posterior probability density function for the parameters given the data, *π*(*θ*|*D*_*obs*_). To this end, we use an approximate Bayesian algorithm which follows the same steps as the minimum distance method proposed by Perez-Reche et al. (*43*); the only difference being that here we use a likelihood function to quantify the similarity of simulated and observed time series instead of a quadratic distance.

The posterior *π*(*θ*|*D*_*obs*_) is approximated by the empirical distribution of a set of 500 point estimates 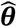 of the model parameters. A point estimate 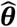 is obtained by simulating *n*_*e*_ = 3000 epidemics with parameters sampled from a prior probability density 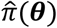. In each realization, a simulation of Model 1 produces deterministic evolution functions *I*_*t*_(*t*) and *D*(*t*) for the number of tested cases and cumulative deaths. The functions *I*_*t*_(*t*) and *D*(*t*) are used to build a random daily time series 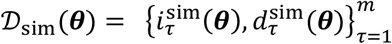, where 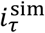 and 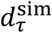 are, respectively, the number of tested infected and deaths predicted at day *τ*. We assume that 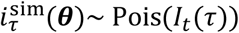 and 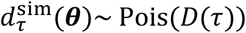, i.e. the predicted number of tested infected and deaths are described as random variables obeying a Poisson distribution with mean *I*_*t*_(*τ*) and *D*(*τ*), respectively. The point estimate 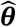 is defined as the parameter vector corresponding to the realization that gives the closest prediction, *𝒟*_sim_, to the observations, *𝒟*_*obs*_. More explicitly, the point estimate for the model parameters is given by

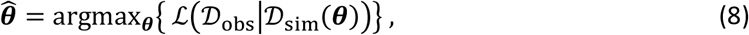

where ℒ(*𝒟*_*obs*_|*𝒟*_sim_(*θ*)) is a log-likelihood function defined as

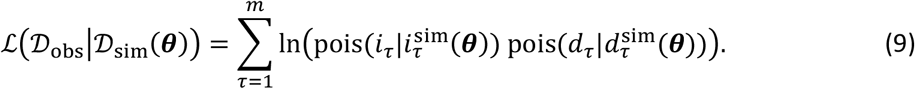

Here,

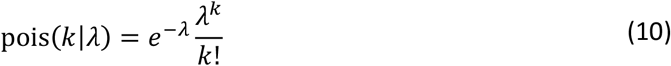

is the Poisson probability mass function.

The prior probability density is defined as the product of priors for each parameter:

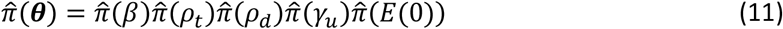

The priors used in our fits are summarized in

Table 3. Normally distributed informative priors were used when prior information on the parameter was available. In particular, since the reproductive number has been more thoroughly studied in previous works than the transmission rate, we set an informative prior for ℛ_0_ and derive *β* from Eq. (4) using the priors of ℛ_0_, *ρ*_*t*_, *γ*_*t*_ and *γ*_*u*_.

**Table 3.** Assumptions for the prior probability distribution of the estimated parameters. 𝒩(*μ, σ*^2^) denotes a normal distribution with mean *μ* and variance *σ*^2^. 𝒰(*a, b*) denotes a uniform distribution in the interval (*a, b*).

| Parameter | Prior | Support |
| --- | --- | --- |
| Fraction tested infected, $\rho_t$ | $\mathcal{U}(0,1)$ | Uninformative for $\rho_t \in [0,1]$ |
| Fraction of tested infected that die, $\rho_d$ | $\mathcal{N}(0.034, 0.01^2)$ | Mean set to the global estimate of WHO (56) |
| Recovery rate for untested infected, $\gamma_t$ | $\mathcal{U}(0,0.4)$ | Range assumed from manual fit exploration (contains typical values for recovery rate (20, 31)) |
| Recovery rate for untested infected, $\gamma_u$ | $\mathcal{U}(0,0.4)$ | Similar assumptions as for $\gamma_t$ |
| Reproduction number, $\mathcal{R}_0$ | $\mathcal{N}(4, 1^2)$ | Estimates from Ref. (4) |
| Transmission rate, $\beta$ | Derived from other parameters using Eq. (4) | |
| Number of exposed at the first data point, $E(0)$ | $\ln E(0) \sim \mathcal{N}(8, 0.5^2)$ | Refs. (12, 20) and manual fit exploration |

## Data Availability

Details on data availability are given in the manuscript.

## Acknowledgments

We acknowledge fruitful discussions with Oliver Carrillo, Bruno Lopes, Christopher McGuigan, Veronica Morales, Stefano Polizzi, Ovidiu Rotariu, John Strachan and Eduard Vives.

